# The use of artificial intelligence and machine learning methods in first trimester pre-eclampsia screening: a systematic review protocol

**DOI:** 10.1101/2022.07.20.22277873

**Authors:** Paula L. Hedley, Christian M. Hagen, Casper Wilstrup, Michael Christiansen

## Abstract

**Introduction:** Pre-eclampsia (PE) is a leading cause of perinatal morbidity and mortality worldwide. Low-dose aspirin can prevent PE in high risk pregnancies if started early. However, despite intense research into the area, first-trimester screening for PE risk is still not a routine part of pregnancy care. Several studies have described the application of artificial intelligence (AI) and machine learning (ML) in risk prediction of PE and its subtypes. A systematic review of available literature is necessary to catalogue the current applications of AI/ML methods in early pregnancy screening for PE, in order to better inform the development of clinically relevant risk prediction algorithms which will enable timely intervention and the development of new treatment strategies. The aim of this systematic review is to identify and assess studies regarding the application of AI/ML methods in first-trimester screening for PE.

**Methods:** A systematic review of peer-reviewed as well as the grey literature cohort or case-control studies will be conducted. Relevant information will be accessed from the following databases; PubMed, Google Scholar, Web of Science, Arxiv, BioRxiv, and MedRxiv. The studies will be evaluated by two reviewers in a parallel, blind assessment of the literature, a third reviewer will assess any studies in which the first two reviewers did not agree. The free online tool Rayyan, will be used in this literature assessment stage. The Preferred Reporting Items for Systematic Reviews and Meta-Analyses (PRISMA) 2020 checklist will be used to guide the review process and the methods of the studies will be assessed using the Newcastle-Ottawa scale. Narrative synthesis will be conducted for all included studies. Meta-analysis will also be conducted where data quality and availability allow.

**Ethics and dissemination:** The review will not require ethical approval and the findings will be published in a peer-reviewed journal using the PRISMA guidelines.

## Introduction

Estimated to cause 100,000 maternal deaths annually [1]; pre-eclampsia (PE) is an frequent (3-8 %) [2] and important contributor to maternal [3, 4] and perinatal [5] — mortality and morbidity. PE is also associated with cardiovascular morbidity and metabolic syndrome later in life for both mother and child [6]. Low dose aspirin treatment reduces the risk of PE, if initiated prior to week 16 [7]. It is therefore important that high-risk patients are identified early, offered preventive treatment, and then carefully monitored throughout pregnancy [5].

Demographic and clinical risk factors along with a plethora of first trimester biochemical and ultrasound markers of the development of PE [8-15] have been identified. These factors and markers have been combined in several recommended algorithms [16] with variable performance [17], particularly for late-onset PE. There is thus a need to develop more effective screening methods [18]. An immense body of knowledge has been assembled on markers and potential risk algorithms and it has been suggested that future developments should rely on large-scale prospective studies [19]. However, recent improvements in computing power and cloud storage will make it possible to use clinical phenotype data from patient registers [20] and novel methods of data combination [21] to develop high-performance, robust, clinical screening algorithms for PE [22]. Using big data in this way may also enable the definition of algorithms that can identify pregnancies which will benefit from preventive treatment, as well as support research into the development and implementation of individualised preventive treatments [23].

Artificial intelligence (AI) and machine learning (ML) methods may provide the key to unlocking the potential of the many markers and risk factors of PE identified to date. By enabling the combination of clinical phenotype information (extracted from electronic health records) and biomarker information along with environmental exposures, AI/ML methods provide the promise of producing a clinically relevant PE prediction algorithm [20]. Furthermore, AI/ML methods have been applied to the prediction of pregnancy complications, albeit not in a clinical care setting [24].

The objective of this systematic review is to identify and assess studies regarding the application of AI/ML methods in first-trimester screening for PE. The research questions (below) follow a PIO method in which the population of interest (P) are first-trimester pregnancies, the intervention (method of analysis) (I) is any AI/ML method, and the outcome (O) is prediction of PE risk.

### Research questions

- Which AI/ML methods (I) have been used to assess PE risk (O) during the first-trimester (P)?
- How effective are these algorithms (I) at predicting PE risk (O) during the first trimester (P)?
- What risk factors have been associated with PE risk (O) during first-trimester (P) using AI/ML methods (I)?

## Methods

In accordance with the guidelines, our systematic review protocol has been submitted to the International Prospective Register of Systematic Reviews (PROSPERO) and is, at the time of submission, awaiting approval.

### Study design

A systematic review of both peer-reviewed and grey literature with a meta-analysis (if possible), will be performed in accordance with the Preferred Report Items for Systematic Reviews and Meta-analyses (PRISMA) statement [25]. The PRISMA Protocols (PRISMA-P) checklist [26] was used to prepare this protocol (S1 Table).

### Eligibility criteria and information sources

We will include original studies (cohort or case–control studies) on PE, performed on samples taken during the first trimester or performed on data which is otherwise not specific to the stage of pregnancy (e.g. genetic variants). AI/ML methods must have been used to assess the data. Inclusion and exclusion criteria are listed in Table 1. Electronic searches of literature will be carried out using the following databases: PubMed, Web of Science, Google Scholar, Arxiv, MedRxiv, and BioRxiv. No date limit will be applied to the searches, with the exception of the pre-print, grey literature sources (Arxiv, MedRxiv, and BioRxiv) which will be limited to articles submitted from the 1^st^ of January 2020 in order to reduce duplication with published sources as well as eliminate publication failures.

**Table 1.** Inclusion/exclusion criteria.

|  | Inclusion | Exclusion |
| --- | --- | --- |
| **Population** | • First trimester pregnancy |  |
| **Methods (Intervention)** | • Artificial intelligence<br>• Machine learning<br>• Integrated learning<br>• Neural network<br>• Sequence learning<br>• Deep learning<br>• Network analysis<br>• Boosting gradient<br>• AdaBoost<br>• XGBoost<br>• Symbolic regression<br>• Supervised learning<br>• Unsupervised learning<br>• Random forest<br>• Decision forest<br>• Decision tree<br>• Support vector machine<br>• Reinforcement learning<br>• Bayesian network<br>• Genetic algorithm<br>• Dimensionality reduction<br>• K-nearest neighbour | • Linear regression<br>• Logistic regression |
| **Outcome** | • Pre-eclampsia |  |
| **Language** | • English | • Non-English |
| **Species** | • Human | • Animal studies |
| **Publication Type** | • Original Research | • Case reports/series<br>• Reviews<br>• Commentaries<br>• Editorials<br>• Letter to editors<br>• Expert opinions<br>• Conference abstracts |
| **Study Design** | • Case-control<br>• Cohort study | • Meta-analyses<br>• In vitro studies |

### Search strategy

Search terms to be used are listed in Table 2. Population terms related to the first-trimester will not be used as the time of testing may be unrelated to the stage of pregnancy (e.g. genetic variants, clinical history). As an example; PubMed would be searched using the following search strategy: (“artificial intelligence”[MeSH Terms] OR “machine learning”[MeSH Terms] OR “integrated learning”[Title/Abstract] OR “neural network”[Title/Abstract] OR “sequence learning”[Title/Abstract] OR “deep learning”[Title/Abstract] OR “network analysis”[Title/Abstract] OR “boosting”[Title/Abstract] OR “AdaBoost”[Title/Abstract] OR “XGBoost”[Title/Abstract] OR “symbolic regression”[Title/Abstract] OR “supervised learning”[Title/Abstract] OR “unsupervised learning”[Title/Abstract] OR “Random forest”[Title/Abstract] OR “Decision forest”[Title/Abstract] OR “Decision tree”[Title/Abstract] OR “Support vector machine”[Title/Abstract] OR “Reinforcement learning”[Title/Abstract] OR “Bayesian network”[Title/Abstract] OR “Genetic algorithm”[Title/Abstract] OR “Dimensionality reduction”[Title/Abstract] OR “K-nearest neighbour”[Title/Abstract] OR “K-nearest neighbor”[Title/Abstract]) AND (“preeclampsia”[Title/Abstract] OR “pre-eclampsia”[MeSH Terms]).

**Table 2.** List of search terms to be applied covering the AI/ML intervention terms and PE outcome terms.

|  | Search Terms |
| --- | --- |
| **Intervention Terms** | "artificial intelligence" OR "machine learning" OR "integrated learning" OR "neural network" OR "sequence learning" OR "deep learning" OR "network analysis" OR "boosting" OR "AdaBoost" OR "XGBoost" OR "symbolic regression" OR "supervised learning" OR "unsupervised learning" OR "Random forest" OR "Decision forest" OR "Decision tree" OR "Support vector machine" OR "Reinforcement learning" OR "Bayesian network" OR "Genetic algorithm" OR "Dimensionality reduction" OR "K-nearest neighbour" OR "K-nearest neighbor") |
| AND |  |
| **Outcome Terms** | "preeclampsia" OR "pre-eclampsia" |

### Study records

#### Data management and selection process

Literature search results will be collected and merged using Endnote (vers 20.3) and, in order to facilitate collaboration among reviewers during the study selection process, transferred to the free online tool Rayyan [27], where duplicates will be removed, and keyword lists based on the inclusion and exclusion criteria will be made in order to facilitate both level 1 and 2 assessment of the literature search results. Prior to the formal screening process, a calibration exercise will be undertaken to pilot and refine the keyword lists. Two reviewers (PLH and MC) will independently assess titles and abstracts against the inclusion and exclusion criteria, as guided by the keyword lists, in order to identify eligible articles. Articles will be rated as ‘included’, ‘excluded’ or ‘maybe’ (i.e. insufficient information in abstract to decide eligibility) and a full-text review will then be performed by both reviewers for all ‘included’ and ‘maybe’ articles. Any discrepancies between the first and second reviewer will be discussed and in the event that a consensus cannot be reached a third reviewer (CMH or CW) will make the final decision regarding eligibility. A PRISMA flow diagram will be drawn to demonstrate the stages of the literature selection process and record the reason for excluding studies (Fig 1) [25].

**Fig 1.**
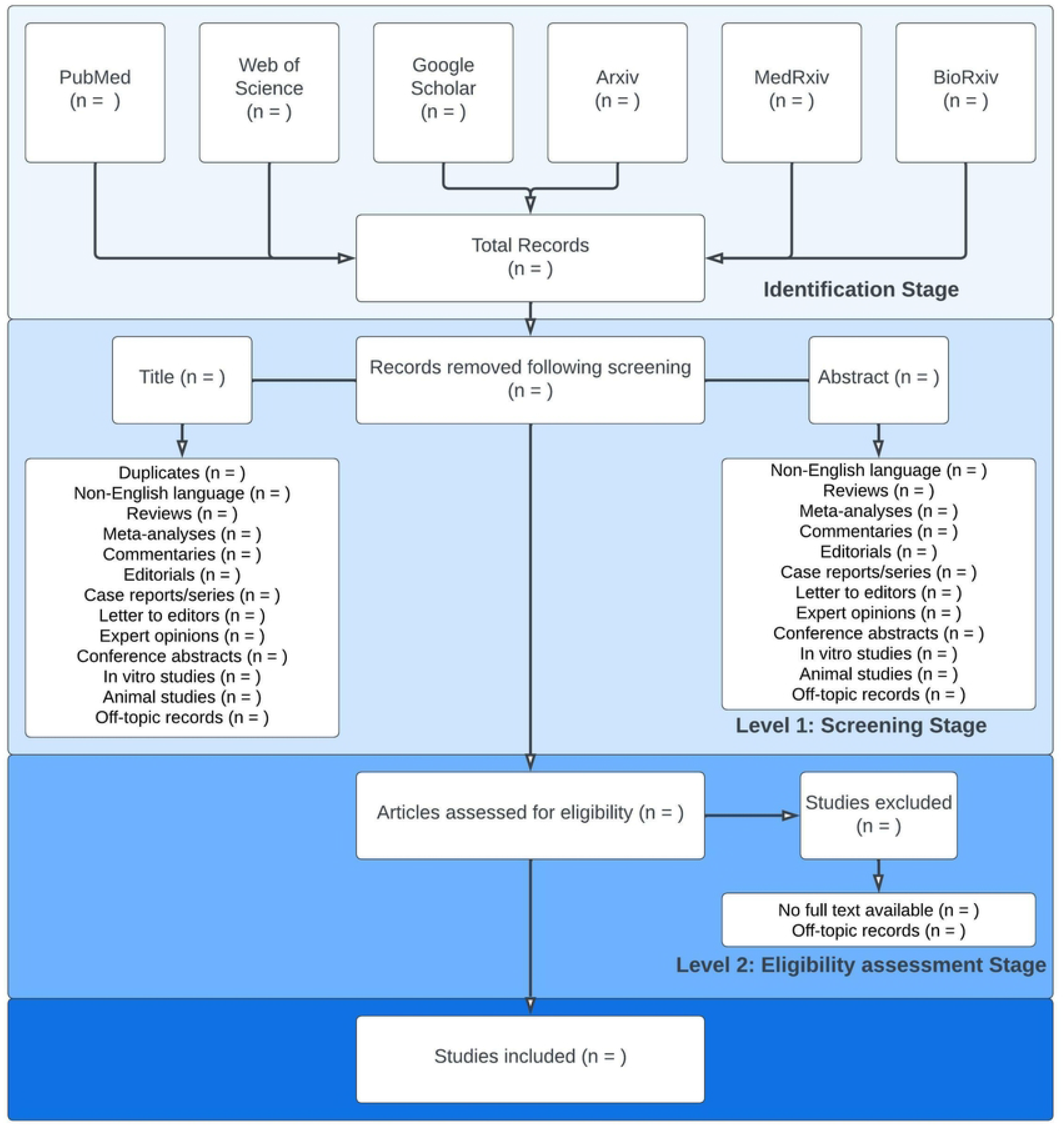
PRISMA Flow diagram. An illustration of the process of selecting studies for review.

### Critical Appraisal of selected studies

The reviewers (PLH and MC) will assess the quality of all eligible studies using the Newcastle-Ottawa Quality Assessment Scale (NOS) for case-control and cohort studies [28]. NOS is a validated quality assessment tool which is divided into three sections focused on selection of participants, comparability of study groups, and ascertainment of outcomes. Each section contains a number of criteria for which a study can be awarded a star, the maximum number of stars that can be awarded is nine. Any discrepancy in NOS scoring will be settled by a third reviewer (CMH or CW). If a discrepancy persists then the average score will be used. Studies with 7-9 stars will be considered high-quality and their data used in a meta-analysis.

### Data collection

One reviewer (PLH) will extract data from the included studies using an excel spreadsheet. Data from multiple reports pertaining to the same study will be linked so as not to report duplicate results. The excel spreadsheet will collect data on the first author, publication year, study period, country of study, type of study, peer-reviewed status, pre-publication status, sample size, maternal age, gestational age at sampling, blood pressure measurements, maternal body mass index, birth weight, risk markers, AI/ML method, all reported outcomes pertaining to PE, and NOS scores. The excel sheet will be adapted following a pilot test on 10 articles, selected to represent various AI/ML methods and risk factors, in order to ensure all relevant data is extracted. The second reviewer (MC) will approve the excel sheet and check the accuracy of the data.

### Data synthesis

Tables will be created to summarise the characteristics of the included studies. A narrative analysis will be presented with reference to the particular AI/ML method and type of data used in the study. We intend to compare the diagnostic efficiency of the different methods, if sufficient studies will be found. If possible, considering the wide range of possible AI/ML methods employed, a meta-analysis will be performed using R package meta. In the event that a meta-analysis cannot be performed, data for similar AI/ML methods will be compiled and reported together. Strength of evidence will be graded by two reviewers (PLH and MC) using the Evidence-based Practice Centre 2015 guidelines [29].

### Patient and public involvement

Patients or the public will not be involved in this systematic review.

### Ethics and dissemination

This systematic review will not need ethical approval because it will retrieve and synthesise data from publically available published and pre-published studies. Study results will be disseminated through scientific publications presented at relevant local and international scientific conferences.

## Discussion

Studies have been performed which use AI/ML methods to predict complications in pregnancy [24]. However, as most studies focus on second trimester pregnancies an overview of first trimester pregnancy PE prediction using AI/ML methods is needed as identification of PE risk in the first trimester can be used to select pregnancies that may benefit from preventive treatment with Low dose aspirin, which has been documented to reduce the occurrence of PE [7, 30].

Many AI/ML methods are currently in use within research and development [21]; this review would allow us to comment on the ability of these methods to support patient participation in decisions with reference to the “explainability” or “interpretability” of the result. These two concepts refer to the ability to explain how the algorithm parameters relates to the result (explainability of black-box models) and how understandable and trustworthy the algorithm results are (interpretability of white-box models) [31]. As risk assessment for PE supports high-risk decisions it is a very important that patients and doctors can understand which underlying factors determine the outcome.

This systematic review will establish the current state of knowledge concerning the prediction of PE during the first trimester through use of AI/ML methods and will provide the highest level of evidence to inform future research as well as the development of first-trimester, PE screening algorithms. However, the considerable heterogeneity expected among the included literature (i.e. different AI/ML methods used, different sample type or gestational ages at time of sampling) may pose a challenge for consistent and comprehensive data extraction. Consequently, meta-analysis will be limited by the quality and quantity of data available. Any significant amendments to this systematic review protocol will require the approval of all authors and will be documented, with the date of the amendment and the rationale for the amendment, on the PROSPERO record, updated protocol document, and in the final manuscript. The review findings will be published in a peer-reviewed open-access scientific journal at the conclusion of this study.

## Data Availability

No datasets will be generated during the current study. All relevant data from this study will be made available upon study completion.

## Supporting information

**S1 Table. PRISMA-P 2015 checklist**

## Statements

### Author contributions

MC is the guarantor. PLH and MC drafted the manuscript and contributed to the development of the selection criteria, the critical appraisal strategy and data extraction criteria. PLH developed the search strategy. CW and CMH provided statistical expertise. CW provided expertise on artificial intelligence and machine learning. All authors read, provided feedback and approved the final manuscript.

## Abbreviations

AI: artificial intelligence
ML: machine learning
NOS: Newcastle-Ottawa Quality Assessment Scale
PE: pre-eclampsia
PRISMA: preferred reporting items for systematic reviews and meta-analyses
PRISMA-P: PRISMA-protocols
PROSPERO: the international prospective register of systematic reviews

